# People who identify as trans, non-binary or gender diverse can experience inclusive, culturally sensitive physical therapy while attending a gender-affirming clinic: qualitative study

**DOI:** 10.1101/2023.10.10.23296844

**Authors:** Madelaine Aird, Julie L Walters, Alex Ker, Megan H Ross

## Abstract

**Objective:** To explore physical therapy experiences and identify barriers and facilitators of access to physical therapy for people who identify as trans, non-binary, or gender diverse (TGD).

**Methods:** A qualitative descriptive design was employed using semi-structured interviews conducted in New Zealand. Participants were individuals aged 12 years or older, who self-identified as TGD, and had accessed physical therapy at a New Zealand clinic that provided a gender-affirming service. Participants were recruited via email invitation to the clinic database. Interview data were analysed using reflexive thematic analysis.

Demographics are reported descriptively.

**Results:** Seventeen individuals participated. Four themes identified during analysis. Barriers to and facilitators of care were identified at policy, environmental, clinic and therapist levels. All participants reported physical therapy experiences relating to one or more of the following themes: *challenging cisnormativity* at policy, environmental, clinic and therapist levels; *safety and trust* throughout the clinical experience-including clinic credibility for being a safe provider, clinic displays of TGD-inclusivity, implementation of safe clinic processes, and respectful therapist interactions; *inclusive experiences* in a clinic which provided affordable care and took active steps to understand and affirm TGD identities, and physical therapists with a high level of knowledge of TGD-specific health issues and a biopsychosocial approach to care; and sensitivity to *body discomfort or dysphoria* triggers.

**Conclusions:** People who identify as TGD face challenges to accessing safe and culturally sensitive physical therapy. However, there are achievable areas for improvement at policy, environmental, clinic and physical therapist level to gain trust and engagement in care for the TGD-community.

**Impact Statement:** This study provides nuanced exploration of TGD physical therapy experiences and identifies specific areas of improvement for TGD physical therapy care to provide clinicians and physical therapy clinics insights into the provision of safe and culturally sensitive physical therapy.

## INTRODUCTION

With increased visibility of people who identify as trans, non-binary or gender diverse (TGD) in Australia and New Zealand (NZ),^1, 2^ the need for TGD-specific healthcare is rising.^3^ In NZ, 2.5% of youths question their gender identity^4^ and 2.3% of Australians aged 15-17 identify as TGD.^5^ Despite increased recognition of gender diversity in public and political discourse over recent years, including legal protections from gender-based discrimination,^6, 7^ significant stigma and inequality remain.^8^ Health, and access to healthcare, are persistent areas of inequity. Individuals who identify as TGD experience poorer physical and mental health than the general population,^9, 10^ including a greater burden of disability, harassment, violence, suicide, suicidality, and homelessness.^11–13^

Globally, health inequities are recognised to emanate from social determinants of health,^14^ the economic and social factors that influence differences in health status. For individuals with TGD identities this includes safe housing,^15^ income,^16^ schooling,^17^ marginalisation,^18^ stigmatisation, and access to and quality of healthcare.^19, 20^ Such determinants can be organized using the socioecological model of health (SEM),^21^ a framework for understanding interconnections between interpersonal, organisational, environmental and policy level factors which impact health. Thus, targeted improvements can be made at each level.

Some, not all, TGD individuals experience gender dysphoria: distress associated with the dissonance between a person’s gender and body.^22^ Exposing the body, observation, and physical touch are routine in the context of physical therapy,^23^ yet TGD experiences of this are largely unresearched. Physical therapy experiences for the broader lesbian, gay, bisexual, transgender, intersex, queer, and related communities (LGBTIQ+) include assumptions of gender, discomfort about touch and undressing, real and perceived discrimination, and lack of trans-specific health knowledge.^23^ However, only ten percent of participants in the study identified as TGD, therefore, in-depth and TGD-specific investigation is required to understand the unique and nuanced factors that influence physical therapy care for this population.

Physical therapists in Australia indicate lacking sufficient knowledge to confidently and sensitively interact with people who identify as TGD identities^24^ and physical therapy students report feeling unprepared and desire further training.^25, 26^ Understanding how people who identify as TGD experience physical therapy and what impacts access to care for this community will provide insights to improve culturally responsive physical therapy for individuals with TGD identities.

Therefore, the research questions for this qualitative study were:

1. What are the experiences of people who identify as TGD with physical therapy in New Zealand?
2. What are the barriers and facilitators to accessing physical therapy for people who identify as TGD in New Zealand?

## METHODS

### Design

A qualitative descriptive design was used. Clients who attended Willis Street Physical therapy (WSP), a musculoskeletal clinic in Wellington, NZ, which provides a gender-affirming (GA) service, and self-identified as TGD were invited to participate. Data were collected using online videoconferencing (Zoom; https://zoom.us/) and analysed with reflexive thematic analysis.^27^ Participants were recruited via email to the clinic’s client database and study information was provided. Participant contact information and demographic data were collected via Qualtrics (www.qualtrics.com). Informed consent of participants was obtained.

Semi-structured interviews were selected to realise the study’s aim of garnering detailed and nuanced descriptions of experiences and the meanings attributed to them.^28^ A purpose-built semi-structured interview guide (see **Appendix 1**) was developed to allow exploration of topics, whilst also allowing discovery of new concepts.^29^ Interview recordings were transcribed verbatim, and pseudonyms ascribed to preserve participant anonymity. A NZ$20 fuel voucher was provided to participants as an honorarium for their time. The funders played no role in the design, conduct, or reporting of this study.

### Participants

Eligible participants had accessed physical therapy services (either GA or general) at WSP, were aged 12 years or older, self-identified or previously self-identified as TGD, and were willing to participate in videoconferencing interviews. Participants were recruited until iterative analyses during data collection displayed adequate depth and repetition of concepts to substantiate the rigor of results.^30^

### Theoretical underpinnings

This study was underpinned by the theory of relativism (i.e., all perspectives are individual and valid), such that people with TGD identities were understood to have different experiences and, thus, there is no singular reality.^29^ The theory of intersectionality (i.e., individual experiences are affected by various factors) also underpinned this project, whereby, physical therapy experiences were interpreted to be affected by an intersection of factors, such as culture, ethnicity, or social class, not solely gender identity.^31^ This generated an approach that affirmed and explored diversity of experiences.

### Data analysis

Reflexive thematic analysis of qualitative data was guided by the six iterative stages described by Braun and Clarke.^32^ Themes and subthemes were generated inductively (i.e., from within the data, not a pre-existing framework) allowing investigative and illustrative interpretation.^33^ Throughout, the researchers made conscious efforts to reflexively question their own preconceptions and assumptions in interpretation. Analyses were conducted by MA (a physical therapy student who undertook qualitative training), and MHR and JLW (qualified physical therapists who identify as LGBTQIA+ and are experienced qualitative researchers).

First, MA read the full dataset and generated preliminary codes. Next, the research team discussed potential themes, to which MA categorised and grouped preliminary codes. Then, the research team discussed and refined themes and sub-themes, and MA re-read the entire dataset, coding any additional data. Lastly, the researchers solidified themes and subthemes and coding was finalised. Subthemes were then mapped to an adapted socioecological model of health (SEM). The analysis was further reviewed by a member of the NZ transgender community to ensure sensitivity to, and inclusivity of the NZ context. Any inconsistencies were discussed and either resolved or included in reporting.

Study rigor was guided by Tracy’s eight key markers for qualitative research quality^34^ and the Consolidated Criteria for Reporting of Qualitative Research (COREQ).^35^ Demographic data were analysed using descriptive statistics.

## RESULTS

### Participants

In total, 84 people responded to the screening questionnaire, with 17 meeting the eligibility criteria and completing interviews. Participants were between 15 and 64 years of age (median 25) and identified with 11 different genders. More than three-quarters (76%) used they/them pronouns. The majority (88%) identified with a gender different from sex assigned at birth, none identified with an intersex experience (i.e., born with a variation of sex characteristics). Most were of NZ European ethnicity (88%), lived in metropolitan/urban areas (88%) and were employed full-time (47%) or students (24%). The sample varied regarding living situation and education level. Details of participant characteristics are provided in **Table 1**. Descriptions of gender identities are given in **Appendix 2**.

**Table 1.** Characteristics of participants.

| Characteristic | Participants (n = 17) |
| --- | --- |
| Age (yrs.), mean (SD) range | 27 (10.9) 15 to 64 |
| **Gender identity, n (%)<sup>a</sup>** |  |
| Non-binary | 11 (65%) |
| Transgender | 8 (47%) |
| Transmasculine | 5 (29%) |
| Transfeminine | 3 (17%) |
| Genderqueer | 3 (17%) |
| Male | 3 (17%) |
| Agender | 2 (12%) |
| Female | 2 (12%) |
| Takatāpui (Māori Rainbow person) | 1 (6%) |
| Genderfluid | 1 (6%) |
| Prefer to self-describe: Queer | 1 (6%) |
| **Pronouns, n (%)** |  |
| They/them/theirs | 9 (53%) |
| He/him | 3 (17%) |
| He/him, they/them | 2 (12%) |
| They/Them/ia | 1 (6%) |
| She, they | 1 (6%) |
| She/Her | 1 (6%) |
| **Place of residence, n (%)** |  |
| Metropolitan/Urban | 15 (88%) |
| Regional | 2 (12%) |
| **Ethnic group(s), n (%)<sup>a</sup>** |  |
| New Zealand European/Pākehā | 15 (88%) |
| Australian | 1 (6%) |
| Māori | 1 (6%) |
| European | 1 (6%) |
| Other | 2 (12%) |
| **Living situation, n (%)** |  |
| With housemates/friends | 9 (53%) |
| With partner | 3 (17%) |
| Alone | 2 (13%) |
| Other | 3 (17%) |
| **Highest level of education, n (%)** |  |
| University | 6 (35%) |
| Post-graduate | 5 (30%) |
| High school | 5 (29%) |
| Primary school | 1 (6%) |

a = Participants able to select multiple options

### Thematic analysis

Four themes were identified during inductive analyses: *challenging cisnormativity, safety and trust, inclusive experiences,* and *body discomfort or dysphoria* (see **Table 2**). Participants’ perspectives and beliefs about encounters (both positive and negative) and suggestions for improvements were spread across themes. Analysis indicates that participants experienced numerous interpersonal (i.e., therapist-patient), organisational (i.e., clinic), environmental (i.e., context and cultural norms), and policy (i.e., government recognition of gender) factors that influence physical therapy experiences and decisions to seek and/or return to care. Individual participant responses are differentiated by assigned pseudonyms.

**Table 2.** Theme descriptions, exemplar quotes and SEM level.

| Theme<br>Subtheme | Description | Exemplar quote | SEM level |
| --- | --- | --- | --- |
| Challenging<br>cishnormativity | Reports of encountering assumptions of gender due to cishnormativity and binary gender norms. | *'I get misgendered so often... you're just coming out so often... it's a small emotional tax to be, "Actually my pronouns are he/him. Or I'm trans".' (Siobhan, transmasculine)* | <u>Policy, environmental, therapist</u>: pervasive societal and cultural norms in relation to the gender binary. |
| Safety and trust | The level of psychological and emotional safety can establish or erode trust in care. |  |  |
| *Clinic credibility* | Establishing clinic credibility for safe care can increase access. | *'... trust is slow to build, as more people talk about it (a service) in their communities, you know it's a safe, approved place to be.'* (John, transgender, transmasculine, genderqueer, male) | <u>Environmental</u>: context of care, referral pathways, and inter-TGD community assurance of safety. |
| *Signposting and clinic processes* | Signals of TGD-inclusivity can facilitate trust but must be accompanied by safe clinic processes. | *'... seeing it (transgender pride flag) there, it's like, "Oh, good. This feels like a place that's for me."' (Jessi, transgender, transfeminine, non-binary), '... it doesn't matter if the physio is culturally competent or good with this kind of stuff if the receptionist isn't. It has to be the whole team.'* (Sita, non-binary) | <u>Clinic</u>: clinic displays of inclusivity that are followed by continuity of inter-clinic processes e.g., pronouns and chosen name use |
| *Respectful therapists* | Respectful interactions with physical therapists can facilitate feelings of safety and trust. | *'... You want to be able to trust that [the physical therapist] won't make weird comments about your body or the way you present'* (Jade, non-binary) | <u>Therapist</u>: professional, non-judgemental, and respectful therapist interactions. |
| Inclusive experiences | Experiences of inclusive, appropriate, and affordable care. |  |  |
| *Subsidised care* | Clinics providing financial subsidies for care influenced decisions to seek care. | *'... whether I could continue [care] would be based on how much I could afford'* (Ellie, transmasculine, non-binary) | <u>Clinic</u>: Clinic services promoting equitable care. |
| *Therapist knowledge of TGD-specific health issues* | Therapist knowledge, understanding and training in TGD-specific health issues. | *'They (physical therapist) understood dysphoria. And I could say, "No, I'm feeling too dysphoric to do that," and they understood what that meant. I didn't have to explain'* (Ashley, transmasculine, non-binary) | <u>Clinic and therapist</u>: educational curricula development, professional training, and individual therapist skills. |
| *Biopsychosocial practice* | Client-led biopsychosocial practice can result in feeling seen as a whole person and empowered in care. | *'In the past, I've felt like I wasn't treated as the expert in my own experience... [at WSP] every appointment feels like a collaboration'* (Rosie, Non-binary, Agender) | <u>Therapist</u>: therapist abdication of control in care can enable client engagement and self-advocacy in care. |
| Body discomfort or dysphoria | Physical therapy may cause experiences of body discomfort or gender dysphoria. | *'Anytime that I'm going to be talking about my body... especially parts of my body that I really dislike, it's always going to be hard.'* (Ellie, transmasculine, non-binary) | <u>Therapist</u>: therapist sensitivity to triggering factors can minimise the impact of body discomfort or dysphoria. |

Abbreviations: SEM; socio-ecological model of health, WSP; Willis Street Physical therapy, a musculoskeletal clinic in Wellington, New Zealand, which provides a gender affirming service, TGD; people who identify as trans, non-binary, or gender diverse identities

#### Theme 1: Challenging cisnormativity

The theme *challenging cisnormativity* encompassed reports of erroneous assumptions or erasure of participants gender identity due to societal norms of cisnormativity and the gender binary. Participants discussed encountering only binary options (i.e., male/female) for sex and/or gender on governmental institution paperwork (e.g., compensation claims) and clinic intake forms. Others experienced perceptions of binary gender in previous physical therapy encounters. For example, Orion (takatāpui/transgender/transmasculine/non-binary/queer) said a physical therapist (not at WSP) talked about their body *‘in terms of male and female [with regard to] exercises, muscle tone, strength-based things…I definitely got misgendered…it was definitely a very gendered view’*. Responses to these experiences were largely consistent.

Participants described feeling exhausted (by repeated misgendering), ‘*invisible’* (Ems, transgender/non-binary/genderqueer/agender), and *‘other[ed]’* (Siobhan). Many expressed frustration that the onus was on them to provide education (e.g., on pronouns).

At WSP, assumptions were largely not encountered, and this was attributed to staff using gender-neutral greetings, non-gendered language (when appropriate), self-describe/multiple gender options, and space for pronouns/chosen name on intake forms. Generally, participants described feeling surprised (at the rarity of the experience), validated and uplifted (because they were recognised) when this happened. Several participants discussed wishing this was the case within physical therapy broadly and suggested therapists should receive cultural sensitivity training that encompasses reflexivity of gender perceptions to address this – *‘…often it’s people’s unexamined views (of gender) that are harmful. Or they hold a view, but they might not know why’* (Sita).

#### Theme 2: Safety and trust

The most common theme, *safety and trust* in care encompassed reports of trusting services and providers to deliver respectful, and psychologically and emotionally safe care. Subthemes were *clinic credibility, signposting and clinic processes,* and *respectful therapists*.

*Clinic credibility*, in relation to safety, was described by many participants as crucial in informing clinic choice. Referrals from trusted services (e.g., GA medical services), affiliations with TGD-friendly community organisations, and TGD-inter-community endorsement was reported as contributing to WSP’s credibility for being a safe clinic. For example, Sita said *‘…I’m always going to ask a queer friend what their experience is before trusting [a service].’* Such endorsements helped participants overcome distrust from previous negative healthcare experiences, as John said *‘‘…it [is] hard to shelve my previous experiences (of transphobia), there’s always [a] level of paranoia and anxiety’*. Some participants also expressed feeling safer (to access care) at WSP because it resided within a *‘safe little Wellington queer bubble’* (Rob, transgender/female) of TGD-acceptance. Participants anticipated this would not be the case in rural or remote settings.

The second subtheme encompassed *signposting and clinic processes* that displayed and ensured TGD-inclusivity at WSP. Signposting included displaying trans and non-binary pride flags, inclusivity statements and website links to other GA health services. Signposting facilitated trust in WSP and promoted care-seeking behaviour, as Jade said, *‘…the fact that there’s little indicators of how you’ll be treated before you have to take that step [to seek care] makes it a lot easier to do.’* Notably, a substantial number of participants expressed concerns for “virtue signalling” and emphasised the importance of safe clinic processes accompanying signposts. Examples of safe clinic processes included signage ‘to use the bathroom that best aligns with your gender’, identifying locations of nearest gender-neutral bathrooms, and continuity of pronoun and correct name use by all staff members and within inter-organisational documentation and data management (preventing ‘deadnaming’). Orion, referencing a past health care encounter, described: *‘…it almost makes it worse when you have people that are really affirming…and then you have someone within the same service that isn’t…it makes it a lot more jarring’*.

The third subtheme involved reports of interactions with *respectful therapists* at WSP that enabled participants to feel safe to be their whole selves within consultations and disclose their TGD-identity (if relevant). In previous physical therapy encounters, many participants described initial apprehension about being judged by physical therapists because of their TGD-identity. For example, John said, *‘…are they [the physical therapist] going to be accepting? Are they okay with it?’*. Several said having to conceal their TGD-identity impacted negatively on their engagement with care – *‘if I can’t be fully open and comfortable about who I am…I’m not going to get the most out of what I need’* (Rosie). Some (particularly younger) participants said they felt uncomfortable disclosing without therapists establishing adequate rapport.

Mostly participants described feeling safe to disclose/discuss TGD-identity with physical therapists at WSP, who were repeatedly described as ‘*professional and friendly’* (Rob). Examples of respectful therapist interactions included stating their own pronouns when introducing themselves, using participants correct pronouns, and addressing gender identity and expression with nuance, which appeared to normalise gender diversity – Siobhan recounted, physical therapists *‘not acting like being trans is weird is always validating…just brining up (in consultations) binding or other aspects of being trans very matter-of-factly, without a change in tone’*.

#### Theme 3: Inclusive experiences

The theme *inclusive experiences* comprised descriptions of inclusive, appropriate, and affordable physical therapy experiences at WSP. The subthemes were *subsidised care, therapist knowledge of TGD-specific health issues* and *biopsychosocial practice*.

*Subsidised care* (for TGD individuals) featured strongly in participant responses and provoked positive and appreciative reactions. Cost of care was described as *‘…so prohibitive’* (Liam, transgender/genderqueer/male) and was a decisive factor in most participants decisions to seek and/or continue care. A small number emphasised this was particularly important given the high “cost” of being TGD (e.g., gender-affirming surgery costs, purchasing new clothes, mental health expenses etc.).

At WSP, participants noted and valued the high standard of *therapist knowledge of TGD-specific health issues*. This included understanding binding, gender dysphoria, fearing not ‘passing’, the importance of gender expression, and basic TGD-terminology. However, this was not always the case. Some participants described experiencing frustration, primarily because they did not feel the WSP physical therapists questions about hormone medications were relevant – Orion wondered, *‘Do they (physical therapists) need to know if I’m on testosterone?’*. One participant described an historical encounter, not at WSP, where a physical therapist dismissed their symptoms – saying their physical therapist *‘…thought, ‘Oh, you’re trans, so how can you have pelvic floor issues?’* (Riley, transgender/transfeminine/female). Many participants said that physical therapists broadly do not have the required level of knowledge to work appropriately with the TGD community – Riley said, in the past *‘I’ve had to do quite a bit of educating (of physical therapists) about the effects of hormones on the body’*. Most participants expressed a desire for increased therapist training on TGD-specific health issues, especially, GA hormone therapy, as it related to muscle bulk.

The last subtheme, *biopsychosocial practice* delivered by WSP physical therapists, was especially prominent in most participants responses. Consultations at WSP were consistently described as *‘client led’* (Jade), and participants felt they were ‘*given a lot of control in [their] own experience’* (Oliver, transgender/male). Participants expressed feeling that their whole selves were seen and ‘*really understood’* (Crystal, non-binary) because multiple aspects of their lives (e.g., mental wellbeing, personal preference, individual circumstances) were considered in treatment and management. For example, Melissa (transfeminine/non-binary/genderfluid) said this approach felt ‘…*integrated into a whole human life’.* Participants reported this engendered their trust in WSP physical therapists, making therapeutic intervention both engaging and more easily implementable into their lives.

#### Theme 4: body discomfort or dysphoria

Finally, several participants discussed *body discomfort or dysphoria* due to touch, exposure, and observation of the body in physical therapy consultations. For example, Siobhan said, a *‘…[physical therapist] watching me closely, [made] me very aware that my body doesn’t look as I’d like it to look and that someone else is also seeing it in a way that doesn’t feel correct to me’*. WSP therapist factors which increased participant comfort were confirming anatomical terminology clients use -*‘…for example, using chest over any feminine words’* (Ellie) and sensitivity to shared gym spaces and mirrors during consultations – *‘coming across an unexpected mirror can trigger some dysphoria’* (John), particularly early in gender-affirmation. Many participants reported physical therapists gaining continual and clear consent for touch was expressly important. For example, Ashley (transmasculine/non-binary) said her WSP physical therapist *‘always asked for consent’* and this was *‘really helpful’.* Other positive factors included sensitivity to draping, consult room privacy, and warning prior to disrobing so participants could mentally and physically prepare (i.e., via clothing choice).

Some also said focusing on therapy outcomes (opposed to the physical body) increased their comfort. A small but notable number of participants discussed past sexual trauma associated with specific body parts/areas and suggested that therapist understanding of and sensitivity to this was valuable.

## DISCUSSION

Our study is the first to investigate access to and experiences of physical therapy for people who identify as TGD. Whilst physical therapy encounters presented challenges to inclusivity for TGD individuals, there were employable strategies to overcome these (**Figure 1**). The key finding of our study is that the most important factor in facilitating physical therapy attendance, and experience quality, is the feeling of safety and inclusion throughout the clinical encounter. Most participants had largely positive encounters within a clinic where specific steps towards TGD-inclusivity were taken. These steps broadly fit into the themes: *challenging cisnormativity, safety and trust, inclusive experiences,* and *body discomfort or dysphoria*. Some negative experiences were also reported, predominantly related to fears around psychological safety of attending physical therapy, rather than WSP itself.

**Figure 1.**
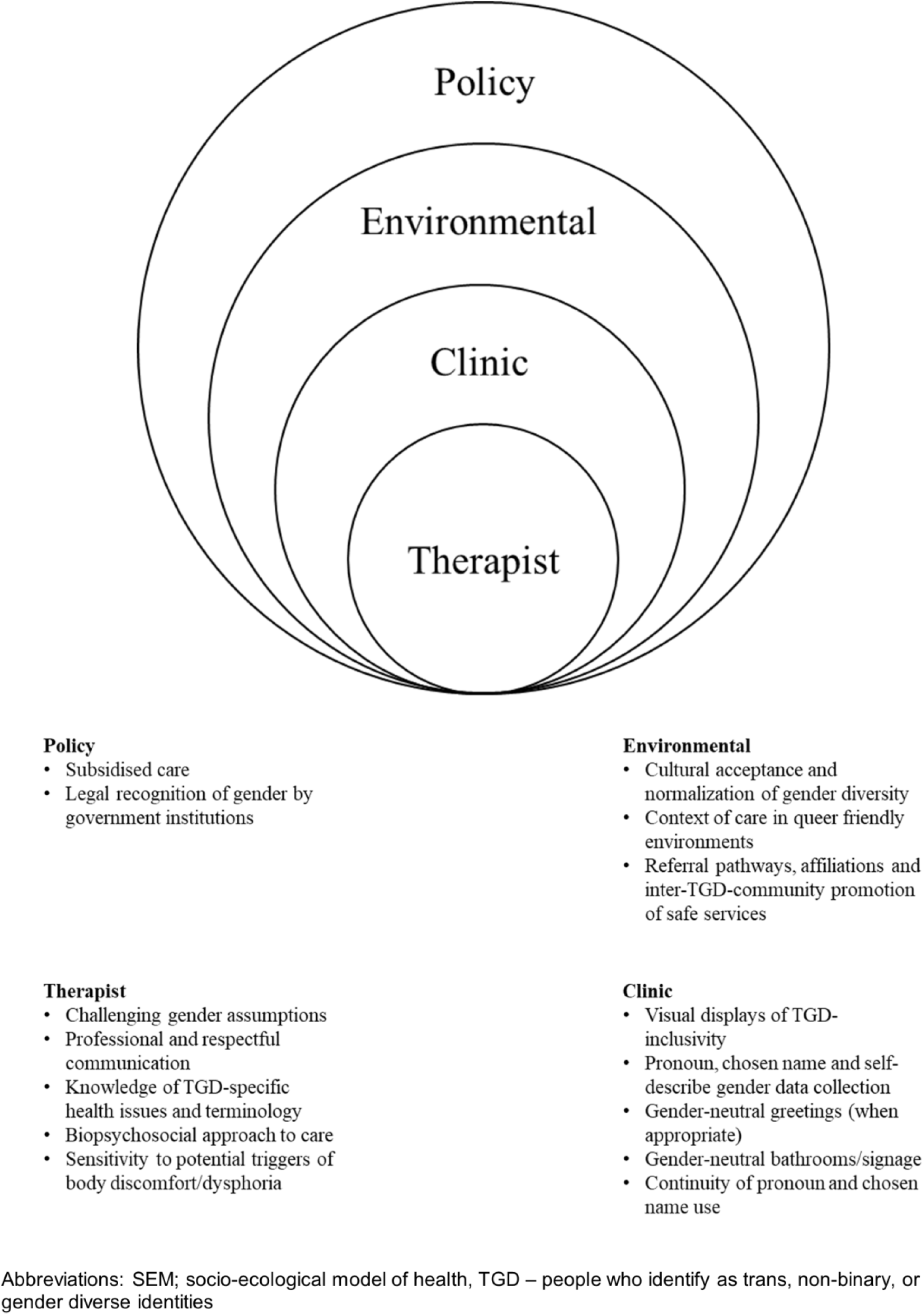
Areas for improvement in physical therapy care delivery at each SEM level.

Our findings were consistent with previous research focusing on the broader LGBTQIA+ community, in that there is pervasive cisnormativity in physical therapy.^23, 24^ As with other healthcare settings,^36–38^ we found misgendering and marginalisation (which may attenuate minority stress^39^ and negatively impact TGD health outcomes^40^) can occur in physical therapy but was noticeably absent at WSP, where our participants attended. Our study found gender recognition at the clinic level crucial to inclusion. Like other findings, participants in our study supported inclusive clinic intake forms (e.g., chosen name, self-describe gender options) and pronoun data collection.^41^

Feeling safe depended upon building trust across all touchpoints of the clinical journey. Most participants expressed fear or anxiety in seeking care because of poor past health care experiences (predominantly within other settings). Real or perceived discrimination is consistently reported as a barrier to TGD access and underutilization of healthcare.^19, 42–45^ Our study identified that safety needs to be established at environmental (i.e., contextual, and social credibility of a clinic for safe care), clinic and therapist levels to facilitate care-seeking and continued engagement in care. Many Australian physical therapists hold largely egalitarian attitudes towards LGBTIQA+ patients.^46^ However, we found that physical therapists may inadvertently misgender and do not always create safe spaces. Challenging therapist cisnormativity through educational training would provide a mechanism for changing physical therapy culture to be more inclusive of gender diversity.

Considering the well-documented stigmatisation and systemic discrimination of this population,^47^ clinics that create safe spaces, in conjunction with physical therapists who upskill in culturally sensitive practice, may help to gain TGD-community trust. The onus is on the physical therapy profession to instigate changes in practice and care delivery. Lack of trust in healthcare encounters is shown to compromise effectiveness of therapy outcomes and can lead to later healthcare avoidance.^48, 49^ Conversely, respectful and trusting therapeutic relationships can improve mental wellbeing for TGD individuals.^50–52^ Furthermore, therapists’ biopsychosocial approach to practice, as described in our study, appeared to gain participants trust. Client-centred care is largely considered gold-standard,^53^ and may promote client self-management^54^ and empowerment.^55^ As lack of trust in healthcare is a persistent barrier for this population,^56^ physical therapists who tailor care to the individual and demonstrate understanding of individual clients’ contexts and socio-emotional needs are likely to facilitate culturally responsive care.

Our findings highlight that affordability was positively associated with physical therapy access. The myriad structural and financial barriers to accessing healthcare faced by the TGD community are well established,^20^ and include socio-economic disadvantage^12^, and employment discrimination^57^ – the unemployment rate of TGD Australians is four-fold the general population^1^ and in NZ, the disposable income of TGD people is one-third less than the cisgender population.^16^ Cost was the final barrier to access for our study’s participants.

Thus, it appears fee subsidisation is a key factor in increasing physical therapy access. For equitable healthcare provision, policy level changes to healthcare costs are required, however, where possible, clinic level measures (e.g., subsidised sessions at WSP) can demonstrate allyship and commitment to equity.

We found high levels of physical therapist training in TGD-specific health issues within WSP, but not in previous physical therapy experiences. In broader healthcare contexts, lack of provider TGD-specific knowledge is frequently reported.^19, 58, 59^ Limited physical therapy-specific TGD-health resources are available, however, research suggests healthcare provider training in TGD care can improve health outcomes and increases knowledge and competence in working with the TGD-community.^60^ Participants in our study supported professional education for basic TGD-terminology and health issues. Further, our findings of body discomfort and dysphoria triggered by physical therapy, provides insight into specific areas of consideration when working with TGD people. Physical therapist sensitivity to factors which could provoke discomfort/dysphoria, and who practiced trauma-informed care, (shown to be beneficial in other healthcare contexts^61^), increased participant comfort where relevant. Whilst there is a lack of physical therapy-specific TGD literature pertaining to physical examination, some non-academic publications (primarily in primary care) exist.^62^ Profession-specific training to increase knowledge of such factors at undergraduate and professional levels offers a way forward to improving care for TGD individuals.

### Limitations

The transferability of our finding requires several considerations. Our study was conducted in NZ which, like Australia, has relatively progressive legislative and social acceptance of TGD people. Findings are likely most applicable to similar socio-economic and political contexts. Our study incorporated a diversity of participants (varied in age and education levels), and findings are likely to be representative of the NZ context. However, sampling within WSP may underrepresent TGD experiences and perspectives of general physical therapy – participants in this study, by majority, only sought physical therapy care at WSP. Despite this, we were able to obtain information about strategies that fostered inclusion and positive physical therapy engagement. Further research into the development and effectiveness of profession-specific TGD-health educational resources would benefit the physical therapy profession.

Our findings highlight that people who identify as TGD can experience inclusive and respectful physical therapy when client-centred care is delivered by clinics and clinicians who have taken active steps towards understanding, including, and affirming TGD-identities and provide affordable care. Access to safe, respectful, and affordable physical therapy would play a role increasing the health status for this population and promote future care engagement. Physical therapists have a moral and professional obligation to provide care which is tailored to the individual and create welcoming and inclusive care for all. Improved awareness and education of TGD-specific barriers to healthcare and health issues offer ways forward for improving TGD physical therapy experiences.

## Data Availability

All data produced in the present study are available upon reasonable request to the authors.

## Ethics approval

The University of Queensland Human Research Ethics Committee (*HE001662)* approved this study. All participants gave written informed consent before data collection began.

## Acknowledgements

The authors would like to thank the participants for their time completing the interviews and questionnaire, Willis Street Physical therapy, Wellington, NZ for their support for this project and assistance with recruitment, and Mr. Alex Ker for his contribution to data analysis.

## Appendix 1. Semi-structured interview guide.

1. Can you please describe your history with physical therapy to date?
  a. Probes/Prompts

i. *How many different clinics, how many different practitioners, over what period of time (in relation to transition / gender identity)*
ii. *What was your main reason for attending?*
2. Did you have any expectations of physical therapy before you went? If so, what were they?
  a. Probes/prompts
    i. *Where did they come from? Why did you think/expect that?*
    ii. *Did it impact your decision about going? (when, where, who)? How?*
    iii. *Did your experience match your expectations? If so, why/how? Why not?*
3. Can you tell me about any negative experiences you had with physical therapy?
  a. Probes/Prompts
    i. *Ask about making appointment, entering clinic, clinic space, staff, consultation, facilities*
    ii. *What made this a negative experience?*
    iii. *How did it make you feel?*
    iv. *Did you go back? Why/why not?*
4. Can you tell me about any positive experiences you had with physical therapy?
  a. Probes/prompts
    i. *Ask about making appointment, entering clinic, clinic space, staff, consultation, facilities*
    ii. *What made this a positive experience?*
    iii. *How did it make you feel?*
    iv. *Did you go back? Why/why not?*
5. Can you describe your experience at Willis Street Physical therapy?
  a. Probes/prompts
    i. *Why/how did you choose this practice?*
    ii. *Was it different to other physical therapy clinics? If so, how?*
    iii. *Were you aware this clinic offers a sex or gender-affirming service? How did you find out? Did that influence your decision, how?*
    iv. *Did you feel safe? Why/how or why/how not?*
    v. *Did you feel the physical therapist had the expertise / knowledge needed for you? Why/why not? Did you like the physical therapist? Why/why not?*
    vi. *Could they have done anything differently? (the clinician and clinic)*
    vii. *Would you refer someone? Why, why not?*
6. What do you wish your physical therapist (or health professionals) knew about the sex and/or gender diverse community?
  a. Probes/prompts
    i. *What do you think is important for physical therapists to know?*
    ii. *Was there anything you have had to teach physical therapists? If so, what?*
7. Is there anything else you would like to add?

## Appendix 2. Glossary of gender terminology

Adapted from the Australian Human Rights commission,^63^ Mental Health Foundation of New Zealand^64^ and Australian Institute of Family Studies.^65^

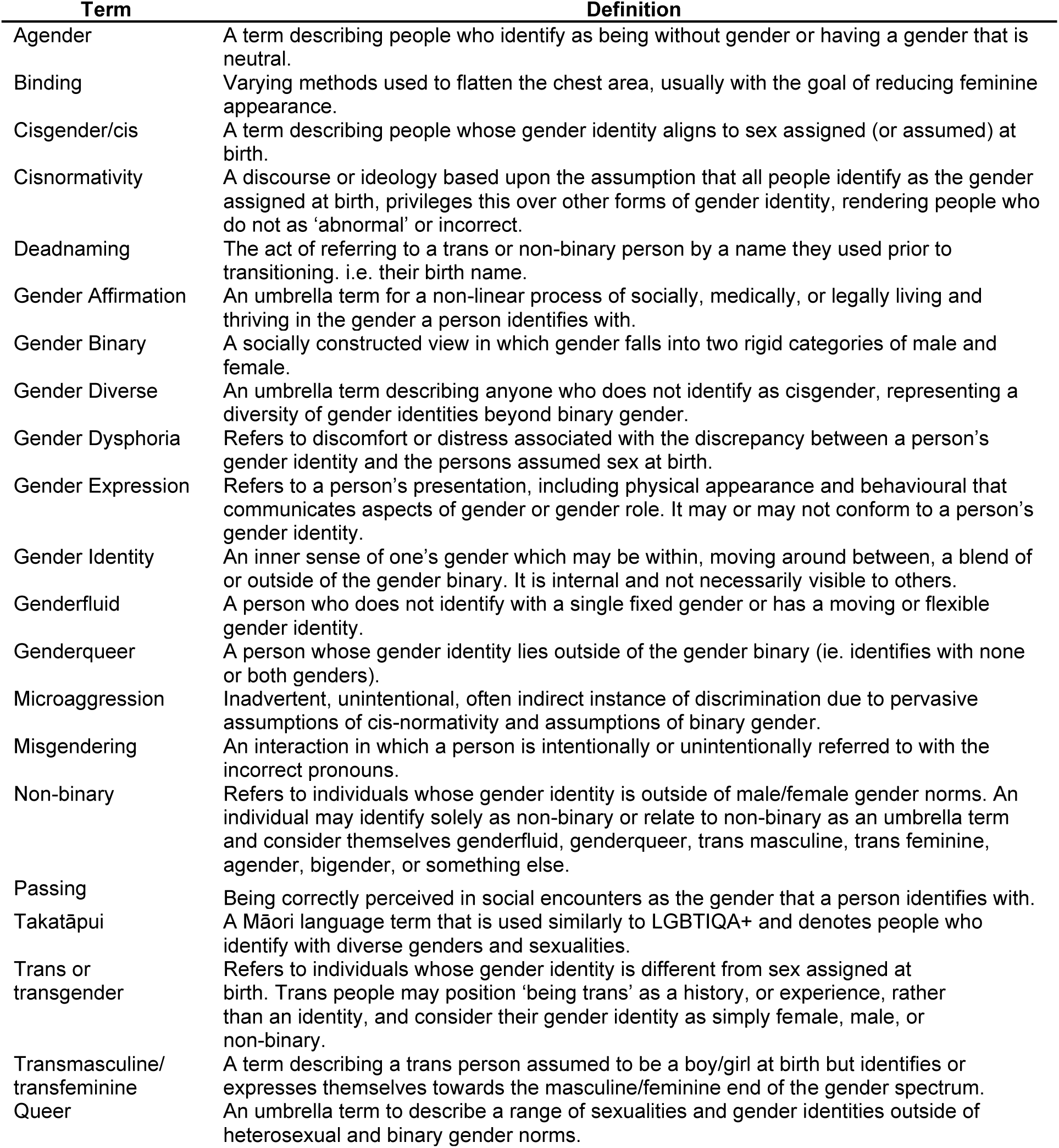

